# Comprehensive analysis of the immunogenomic landscape and clinical features in cervical cancer

**DOI:** 10.1101/2021.08.24.21262578

**Authors:** Xinyue Fan, Chunbo He

## Abstract

Immunotherapy has changed the standard of treatment for many cancers. However, the same treatments showed disappointing outcomes in cervical cancer so far. Thus, understanding the mechanisms that support the immune tolerance of cervical cancer will provide a way to design new strategies to enhance immunotherapies. Here, we characterized cellular compositions of the immune infiltrates in cervical cancer and investigated if the tumor immune landscape is a predictor for patient prognosis. The fraction of ten immune infiltrates of cervical and other cancers were analyzed by using QuanTIseq software base on the bulk mRNA sequencing data from The Cancer Genome Atlas Program (TCGA). Cervical cancer is one of the cancers that had the lowest percentage of total immune infiltrates, but it had the highest ratio for CD8 T cells to all immune infiltrates among all solid cancers. Both the principal components (PCA) analysis and heatmap with dendrogram analysis showed that cervical cancer had a similar immune infiltrated microenvironment with other squamous cell carcinomas, such as head and neck cancer and lung squamous cell cancer. The PCA and heatmap with dendrogram analysis showed that cervical cancer and HPV positive head and neck cancers were clustered more closer and partially separated with HPV negative head and neck cancer. Further analysis showed that HPV-positive cervical and head and neck cancers had a significantly higher level of CD8 T cells and M1-liked macrophages, but a lower level of M2 macrophages. The survival analysis showed that a higher level of CD8 T cells was associated with a better patient prognosis. However, immuno-suppressive immune infiltrates including M2 macrophages and Treg cells that are known to suppress anti-tumor immunity also demonstrated positive patient overall survival. Our study provided a conceptual framework to understand the tumor immune microenvironment of cervical cancer. Our results also demonstrated that the immune infiltrates can be a prognosis marker for cervical cancer.

**Simple Summary:** Cervical cancer is the most common gynecologic cancer and the fourth leading cause of cancer-related death in women worldwide. There are relatively limited treatment options for late-stage cervical cancer. Immunotherapy is a new therapeutic treatment developed with great success in treating many cancers, but the same treatment has not been producing satisfactory results in many cases of cervical cancer. In the present study, we provided a comprehensive immune characterization specifically for cervical cancer. We determined the prognostic value of a specific subtype of tumor-infiltrating immune cells for clinical outcomes and demonstrated that HPV infection affected the immune cell infiltration and induce pro-inflammatory phenotypes. Our study provides a systematic insight into the tumor immune microenvironment of cervical cancers and offers a conceptual framework for the future design of rational combination treatment strategies to improve immunotherapy outcomes.

## 1. Introduction

Cervical cancer is the most common gynecologic cancer and the fourth leading cause of cancer-related death in women worldwide. The International Agency for Research on Cancer (IARC) estimates that there are approximately 527,000 women who are diagnosed with cervical cancer, and more than 265,000 women die of this disease each year [1]. Since cervical cancer is mostly diagnosed between women of the ages 35 to 55, it affects women when they are young and has devastating effects with a very high human, social, and economic cost [2]. Human papillomavirus (HPV) infection is believed to be the major causative factor for cervical cancer, and 99% of cervical tumors are HPV positive. The increased use of early detection and HPV vaccination have significantly reduced cervical cancer-related death in developed countries in past decades [3]. More than 85% of cervical cancer occurred in low- and middle-income areas such as Africa and South Asia [2]. Cervical cancer has a relatively good response towards standard therapies if the cancer is diagnosed in the early stages. However, the prognosis of advanced-stage cervical cancer patients is very low. There are relatively limited treatment options for late-stage cervical cancer when metastasis has occurred or when there is a recurrence of cancer [4]. As a result, new therapeutic treatments need to be developed in order to solve this urgent matter.

Immunotherapy is a new therapeutic treatment developed with great success in treating many types of cancers, including some late-stage and previous terminal illness cancers [5]. For example, the combination of immune checkpoint blockades (ICB) such as anti-PD 1/PD-L1 antibodies and chemotherapy have demonstrated durable response and unparalleled clinical benefit in advanced lung cancer patients [6], CD19 directed Chimeric antigen receptor (CAR) T-cell therapies have demonstrated significant clinical benefit in diffuse large B cell lymphoma (DLBCL)[7]. However, despite these successes, immunotherapy has not been producing satisfactory results in many cases of cervical cancer treatments [8]. Various clinical trials using standard immunotherapy treatment produced poor results for cervical cancer treatments, indicating that the tumor microenvironment of the patient may develop immune resistance and result in poor treatment. Therefore, mechanisms behind this response need to be explored in order to develop better treatment strategies to combat the problems.

The tumor immune microenvironment is demonstrated to play a critical role in cancer initiation, progression, and control [9]. It consists of tumor-associated fibroblast, immune cells, and endothelial cells. The molecular and cellular composition of the tumor microenvironment affects the disease outcome by promoting a suppressing or cytotoxic response around the tumor. Using mRNA sequence data and bioinformatics tools, recent studies characterized the immune microenvironment in pan-cancer or cancer-specific settings and their association with clinical outcomes [10-14]. However, there are few studies that provide a comprehensive immune characterization specifically for cervical cancer.

In the present study, we want to characterize the cellular compositions of the immune infiltrates in cervical cancer. We will try to cluster immune subtypes based on the immune landscape in the tumor microenvironment. We will also analyze the association between immune subtypes and patients’ prognosis in cervical cancer. We hypothesized that a ‘pro-inflammatory’ phenotype, which is when tumors have higher DCs, M1-like macrophages, infiltrating T cells, and Natural Killer cells, was associated with better clinical outcomes. On the other hand, an “immunosuppressive” phenotype that has more inhibitory immune cells like M2-like macrophages, myeloid-derived suppressor cells (MDSCS), and Treg cells were associated with the worse outcome in cervical cancer. This study aims to provide a better understanding of the tumor immune microenvironment and the effect it has on therapeutic responses. This study may also offer a conceptual framework for the future design of treatment strategies to-wards specific groups of patients in order to improve immunotherapy outcomes.

## 2. Materials and Methods

### 2.1. Patient Information and Datasets

The data collection and analysis processing steps were shown in Figure 1. The clinical information of cervical cancer patients was obtained from the Memorial Sloan Kettering Cancer Center on the cBioPortal website (http://www.cbi-oportal.org) [15]. 297 cervical cancer cases were used for analysis. The upper quartile normalized RSEM mRNA gene expression data was obtained from the Broad Institute FireBrowse portal (http://gdac.broadinstitute.org) [16]. Informed patient consent was waived because all data are public data sets that are pre-existing and de-identified. The raw data, processed data, and clinical data came from the legacy archive of NCI’s Genomic Data Commons (GDC) (https://por-tal.gdc.cancer.gov/legacy-archive/search/f). Three genomic subtypes of cervical cancer samples, EMT, Hormone, and PI3K-AKT types, and HPV types were collected from the study Integrated Genomic and Molecular Characterization of Cervical Cancer at Nature [17]. Six immunogenomic subtypes of cervical cancer samples, C1 to C6, were obtained from the study The Immune Land-scape of Cancer at Immunity [10].

**Figure 1:**
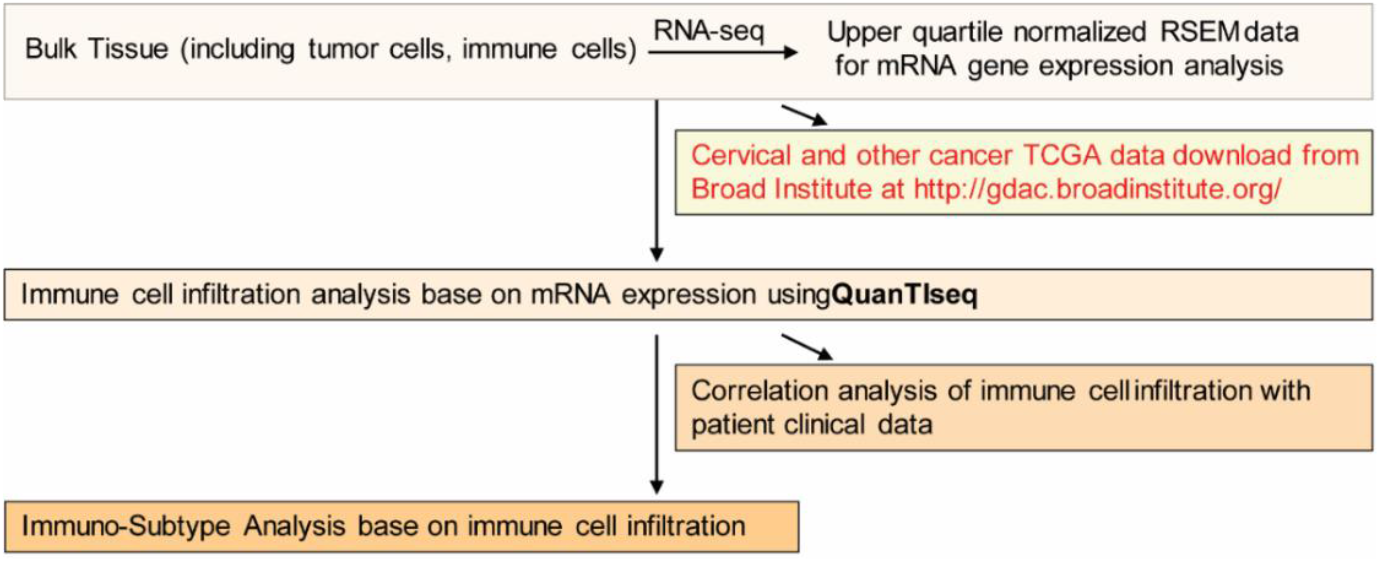
Flowchart of the study process, data collection, and methods used.

### 2.2. Analysis of Tumor-Infiltrating Immune Cells

The composition and density of immune cells in the tumor microenvironment (TME) were analyzed using QuanTIseq, a deconvolution-based method used for quantifying and calculating the fractions of immune cell types from bulk RNA-sequencing data in tumor and blood samples [18]. The QuanTIseq analysis provided the fractions of ten immune cells compared to all cells in the samples selected. The immune cells included are B cells, CD4 T-cells (non-regulatory CD4+ T cells), CD8+ T cells, DCs (dendritic cells), M1 Macrophages (classically activated macrophage), M2 Macrophages (alternatively activated macrophages), Monocytes, Neutrophils, NK cells (natural killer cells), and Treg cells (regulatory T cells). The information of QuanTIseq software pipeline is available at http://icbi.at/quantiseq and https://10.1186/s13073-019-0638-6. In all figures of the present study, we presented QuanTIseq analysis results.

### 2.3. Identification and Visualization of Immuno-subtypes of the Tumor Samples

Immune cell infiltration for cervical cancer was compared with two other gynecological cancers: ovarian cancer and uterine cancer, and with two other squamous cell carcinomas: head and neck cancer and lung cancer. An unsupervised principal component analysis (PCA) and heatmaps with dendrograms were performed on all analyzed samples based on the fraction of ten infiltrating immune cells as determined by QuanTIseq. The analysis was based on the methods reported by Vichi et al. and Granato et al. [19, 20]. Principal Component Analysis (PCA) was visualized through the built-in R functions prcomp().and the visualized first two (PC1 and PC2) projection scatterplots were built using the ggplot2 package of R function.

### 2.4. Correlation of Immune Cell Subtypes with Patient Survival

Survival analysis of cervical cancer patients stratified by the infiltrated immune cell concentration was determined by results from QuanTlseq analysis. For each specific immune cell, survival data with higher (top 50%) immune cell fraction and lower (bottom 50%) immune cell fraction were compared using Kaplan-Meier survival estimates with 95% confidence intervals through using Prism V. 9 (San Diego, CA). P-values were calculated using the Log-Rank test in order to determine whether there were significant differences between the two survival curves for each immune cell infiltration.

### 2.5. Correlation Analysis Between HPV Infection and Immune Cell Infiltration

The immune cell infiltration data of cervical cancer HPV+ patients, Head and Neck Cancer HPV-patients, and Head and Neck Cancer HPV+ patients were obtained from QuanTlseq in order to analyze the differences in immune cells infiltration landscape among the three groups based on the ten immune cell infiltration values. PCA was performed using the built-in R functions prcomp().and the visualized first two (PC1 and PC2) projection scatterplots were built using the ggplot2 package of R function. Box and whisker graphs were also used to analyze the specific types of immune cell infiltrate in HPV+ patients and HPV-patients.

### 2.6. Correlation of Immune Cell Infiltration and genomic and Immunogenomic subtypes

Three genomic subtypes of cervical cancer samples, including EMT, Hormone, and PI3K-AKT types, and six immunogenomic subtypes of cervical cancer samples, C1 to C6, were obtained from the previous studies[10, 17]. T-tests with unequal variances and two-tailed variances were performed to analyze between the C1 and C2 immunogenomic sub-types in terms of immune cell infiltration. One-way ANOVA with Tukey’s post-test was performed to analyze the differences among three TCGA molecular subtype groups (Hormone, PI3K_AKT, EMT) in terms of immune cell infiltration. For both tests, p values < 0.05 were considered as statistically significant.

## 3. Results

### 3.1. The Landscape of Immune Infiltration in Cervical Cancers

To analyze and compare the immune cell subtype composition among different cancer types, the upper quartile normalized RSEM (RNA-Seq by Expectation Maximization) mRNA-seq data from more than 8000 TCGA tumor samples across 19 solid cancer types were collected from the Broad Institute FireBrowse portal (http://gdac.broadinstitute.org). The proportions of 10 types of infiltrating immune cells were distinguished by performing QuanTIseq analysis on these tumor samples based on the mRNA gene expression profile.

As shown in Figure 2A, Glioblastoma (GBM), kidney cancer (KIRC), lung adenocarcinoma (LUAD), and skin cutaneous melanoma (SKCM) contained the highest concentration of immune infiltrates. On the other hand, cervical cancer contained one of the lowest percentages of immune cell infiltrates in solid tumor cells when compared to other types of cancer. Specially, B cells, CD4 T cells, and Dendritic cells. However, when looking at the percentage of CD8 T cells to total immune infiltrates in the solid tumor alone, cervical cancer had the highest percentage of CD8 T cells compared to all 18 other cancers (Figure 2B).

**Figure 2.**
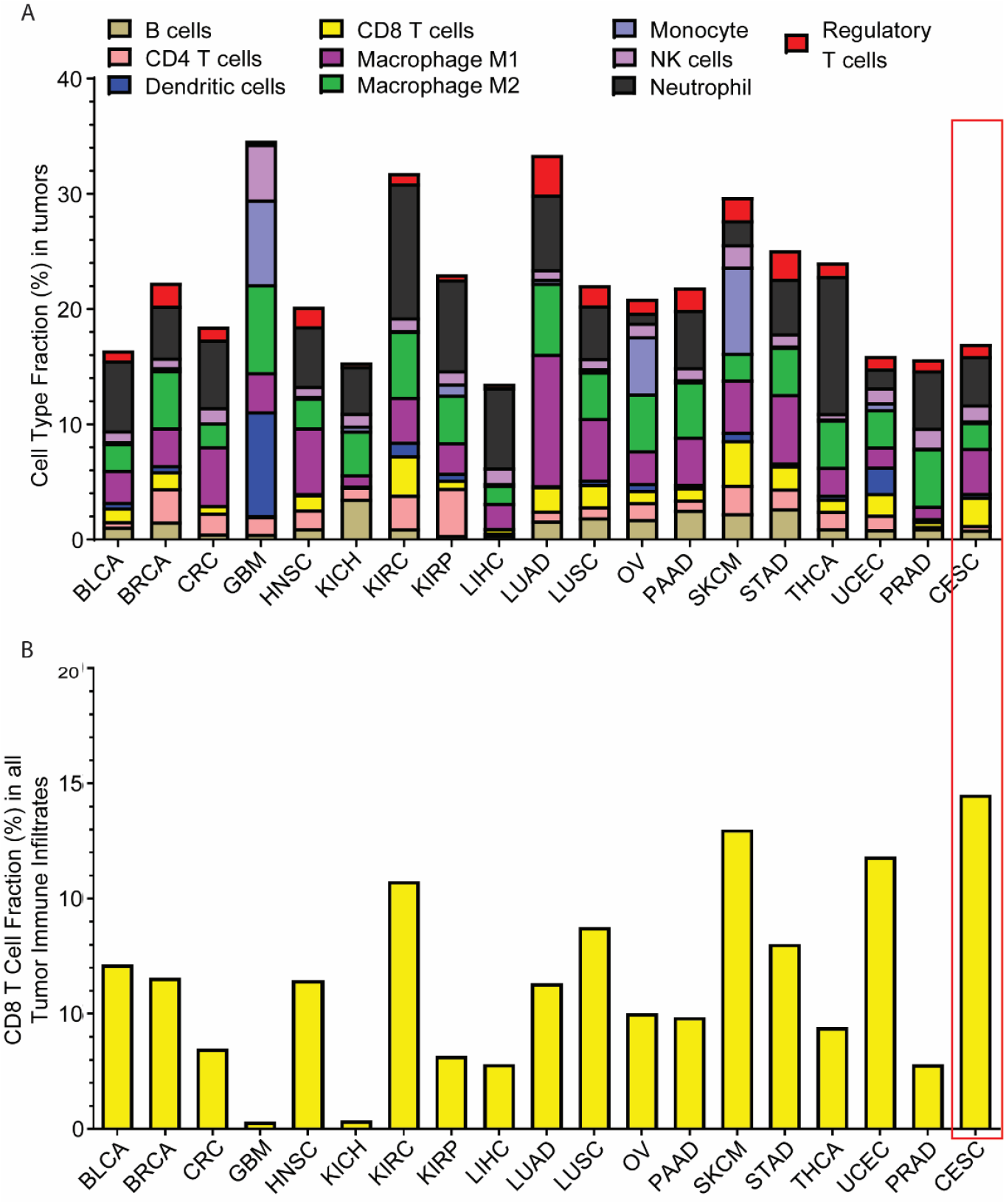
The overall landscape of immune infiltration in cervical cancer. (a) The percentage of each immune infiltrate in Bladder Urothelial Carcinoma (BLCA), Breast invasive carcinoma (BRCA), Colorectal cancer(CRC), Glioblastoma multiforme (GBM), Head and Neck squamous cell carcinoma (HNSC), Kidney Chromophobe (KICH), Kidney renal papillary cell carcinoma (KIRC), Kidney renal papillary cell carcinoma (KIRP), Liver hepatocellular carcinoma (LIHC), Lung adenocarcinoma (LUAD), Lung squamous cell carcinoma (LUSC), Ovarian serous cystadenocarcinoma (OV), Pancreatic adenocarcinoma (PAAD), Skin Cutaneous Melanoma (SKCM), Stomach adenocarcinoma (STAD), Thyroid carcinoma (THCA), Uterine Corpus Endometrial Carcinoma (UCEC), Prostate adenocarcinoma (PRAD), and Cervical squamous cell carcinoma and endocervical adenocarcinoma (CESC). (b) The percentage of CD8 T cells in all immune infiltrates in BLCA, BRCA, CRC, GBM, HNSC, KICH, KIRC, KIRP, LIHC, LUAD, LUSC, OV, PAAD, SKCM, STAD, THCA, UCEC, PRAD, and CESC.

Having relatively lower total immune infiltrates indicates that cervical cancer has a relatively inactive immune environment compared to other types of cancers, especially immunotherapy well-responded cancers such as LUAD and SKCM [6, 21-24]. Cervical cancer also has a high concentration of CD8 T cells compared to 18 other popular cancers. CD8 T cells have a higher contribution to the immune response in current ICB-based immunotherapies [25]. Therefore, treatments that target toward improving T cells functions such as Anti-PD1 and PDL1 may result in better treatment response towards cervical cancer.

### 3.2 Comparison of tumor immune infiltrates among gynecologic cancers and main types of squamous cell cancers

Cervical cancer is one of the most popular types of gynecological cancers and squamous cell cancers. So, we further compared the features of the fraction of infiltrated individual types of immune cells in gynecologic cancers with other main types of squamous cell cancers. Figure 3A showed the absolute proportions of each of the 10 immune cells that were compared among ovarian, cervical, uterine cancers. Cervical cancer had an average higher level of M1 Macrophages and neutrophils, two pro-inflammatory immune cells, compared to uterine and ovarian cancer. Cervical cancer also contains lower levels of M2-Macrophages, an immunosuppressive immune infiltrate compared to two other types of cancers. This data indicates that cervical cancer has an overall more immune-active environment compared to the two other gynecological cancers.

**Figure 3.**
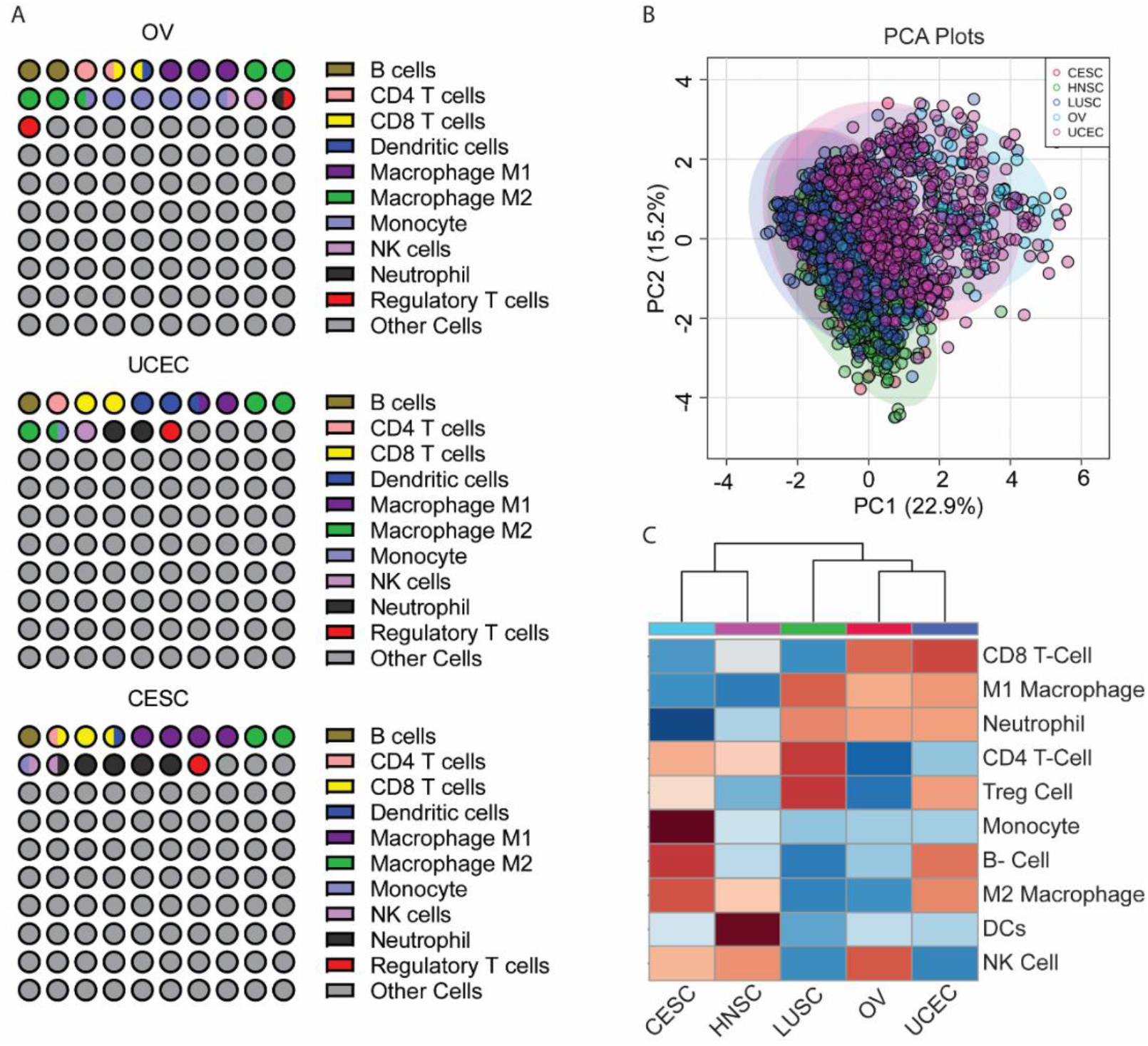
Cervical cancer shared a similar immune infiltrated microenvironment with other squamous cell carcinomas. (A) The percentage of each immune infiltrate in cervical cancer, ovarian cancer, and uterine cancer. (B) & (C) Comparison of tumor immune infiltrates in cervical cancer, head and neck cancer, lung cancer, ovarian cancer, and uterine cancer with principal component analysis and heatmap with dendrograms.

An unsupervised principal component analysis (PCA) and heatmaps with dendrograms were performed to analyze the overall similarity of immune infiltrates landscapes in gynecologic cancers (ovarian, cervical, uterine cancers) and squamous cell cancers (lung squamous cell cancer and head and neck cancer). As shown in Figure 3B & 3C, cervical cancer had a more similar immune cell infiltration microenvironment with squamous cell carcinomas cancer (lung cancer and head and neck cancer) compared to gynecological cancers (uterine cancer and ovarian cancer) due to greater overlap on the PCA plot and closer dendrogram distance between the two groups. This result implies that immune cell-based treatments that are effective towards treating lung cancer and head and neck cancer can also be used for cervical cancer due to their similarity in the immune cell infiltration landscape.

### 3.3 Correlation of Immune Cell Subtypes with Patient Survival in cervical cancer

The association between the tumor infiltrated immune cells and the prognosis of cancer patients had been reported in different types of cancers. For example, a high level of tumor infiltrated CD8 T cells was shown associated with longer survival in melanoma, colon cancer, and lung cancer [12, 26, 27]. To evaluate the prognostic value of tumor-infiltrating immune cells, we analyzed the patients’ overall survival rate relative to the fraction of each immune cell in cervical cancer. As shown in Figure 4, patients that contained higher levels of CD8 T cells are associated with better survival outcomes. This again can serve as an implication that treatments targeted towards greater CD8 T cell efficiency and higher concentrations can result in better and more effective cervical cancer treatment. However, surprisingly, a higher concentration of immune infiltrates that are associated with immuno-suppressive phenotypes such as M2 Macrophages are associated with better survival outcomes. This contradicts our hypothesis because we predicted that a higher concentration of immuno-suppressive phenotype infiltrates is associated with lower survival rates compared to having lower concentrations. The mechanisms behind this result are unknown, a possible reason that can explain is the stroma cell concentration [28, 29]. A higher concentration of M2 Macrophages is usually associated with a higher concentration of stromal cells [30]. Higher concentrations of stroma cells cause a harder time for tumor cells to divide and spread to other areas of the body and therefore may contribute to a higher survival rate.

**Figure 4.**
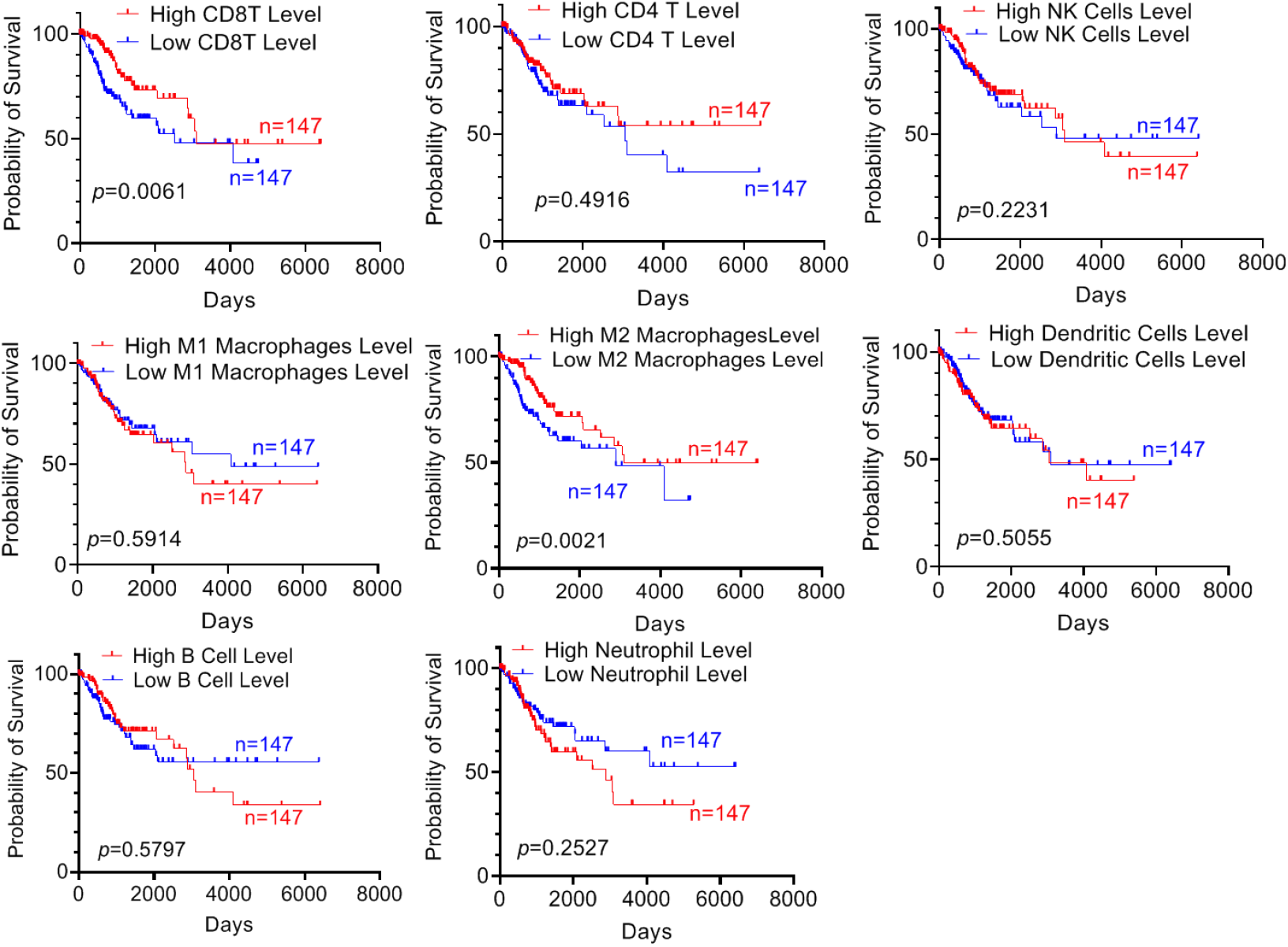
The prognostic value of immune infiltrate immune cells in cervical cancer. The Kaplan-Meier curves are stratified by the proportion of infiltrated immune cells. For each immune cell, survival data with higher (upper 50%) and lower (lower 50%) were compared using Kaplan-Meier survival estimates. P-value was calculated by the log-rank tests. P-values < 0.05 were considered as statistically significant.

### 3.4 Correlation Analysis Between HPV Infection and Immune Cell Infiltration

Infection of high-risk HPV is the main causative factor for cervical cancer, so we also analyzed the HPV infection versus tumor immune cell infiltration. Since more than 99% of cervical cancers are HPV positive [31], we compared HPV-positive cervical cancer patients with Head and Neck cancer HPV-positive and HPV negative patients. Based on the PCA plot and heat graph with dendrogram (Figures 5A & 5B), the immune cell infiltration landscape is similar between HPV positive cervical cancer patients and HPV positive Head and Neck cancer patients due to the greater overlap on the PCA plot and closer dendrogram distance. Further analysis showed that tumors with HPV infection have higher infiltration of pro-inflammatory immune cells like CD8 T cells and M1 Macrophages such as CD8 T cells and M1 macrophages (Figures 5C, 5D, & 5E). These results suggest that HPV infection could strongly affect the immune cell infiltration and induce pro-inflammatory phenotypes. On top of that, the results can also imply that effective treatments currently used for HPV-positive head and neck patients may also be used for cervical cancer patients because of the similarity in immune cell infiltration landscape. These results also indicate the importance of CD8 T cells in cervical cancer and the possibility of developing and using treatments that aim to improve the quantity and efficiency of CD8 T cells in cervical cancer patients.

**Figure 5:**
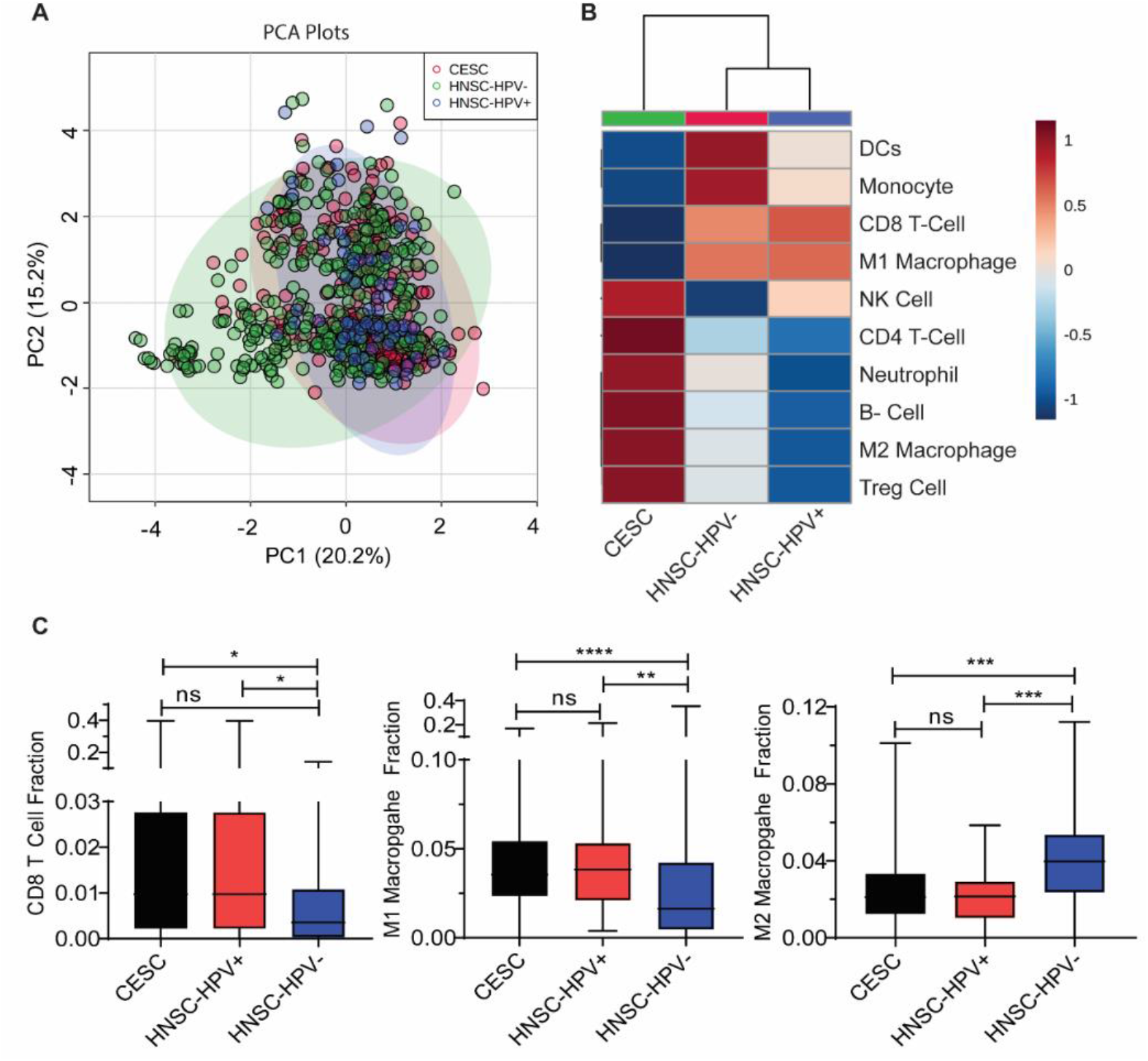
HPV infection affected immune cell infiltration and induce pro-inflammatory phenotypes. (A) & (B) PCA plot and heatmap with dendrogram analysis on the immune cell infiltration landscape comparison between cervical cancer, head and neck cancer HPV positive, and head and neck cancer HPV negative. (C), (D)& (E) Comparison of tumor immune infiltrates among cervical cancer, head, and neck cancer HPV positive, and head and neck cancer HPV negative. One-way ANOVA with Tukey’s post-test was used to analyze differences in fractions of infiltrating immune cells for the three categories. P values < 0.05 are considered statistically significant.

### 3.5 Correlation of Immune Cell Infiltration and genomic and Immunogenomic subtypes

Cervical cancers have substantial heterogeneity. As a result, the prognosis and treatment response are highly variable among patients. A recent study conducted by The Cancer Genome Atlas (TCGA) Research Network for cervical cancer defined three subtypes (Hormone, PI3K_AKT, and EMT subtypes) based on molecular and genetic features of cervical tumors [17]. To investigate if the immune infiltrates are associated with TCGA genetic subtypes, we stratified immune cell fraction according to the three TCGA subtypes. As demonstrated by Figure 6A, based on one-way ANOVA tests, there were no significant differences in terms of immune cell infiltration between each group, indicating that molecular subtypes may not play a strong role in cervical cancer patient’s immune cell infiltration landscape.

**Figure 6.**
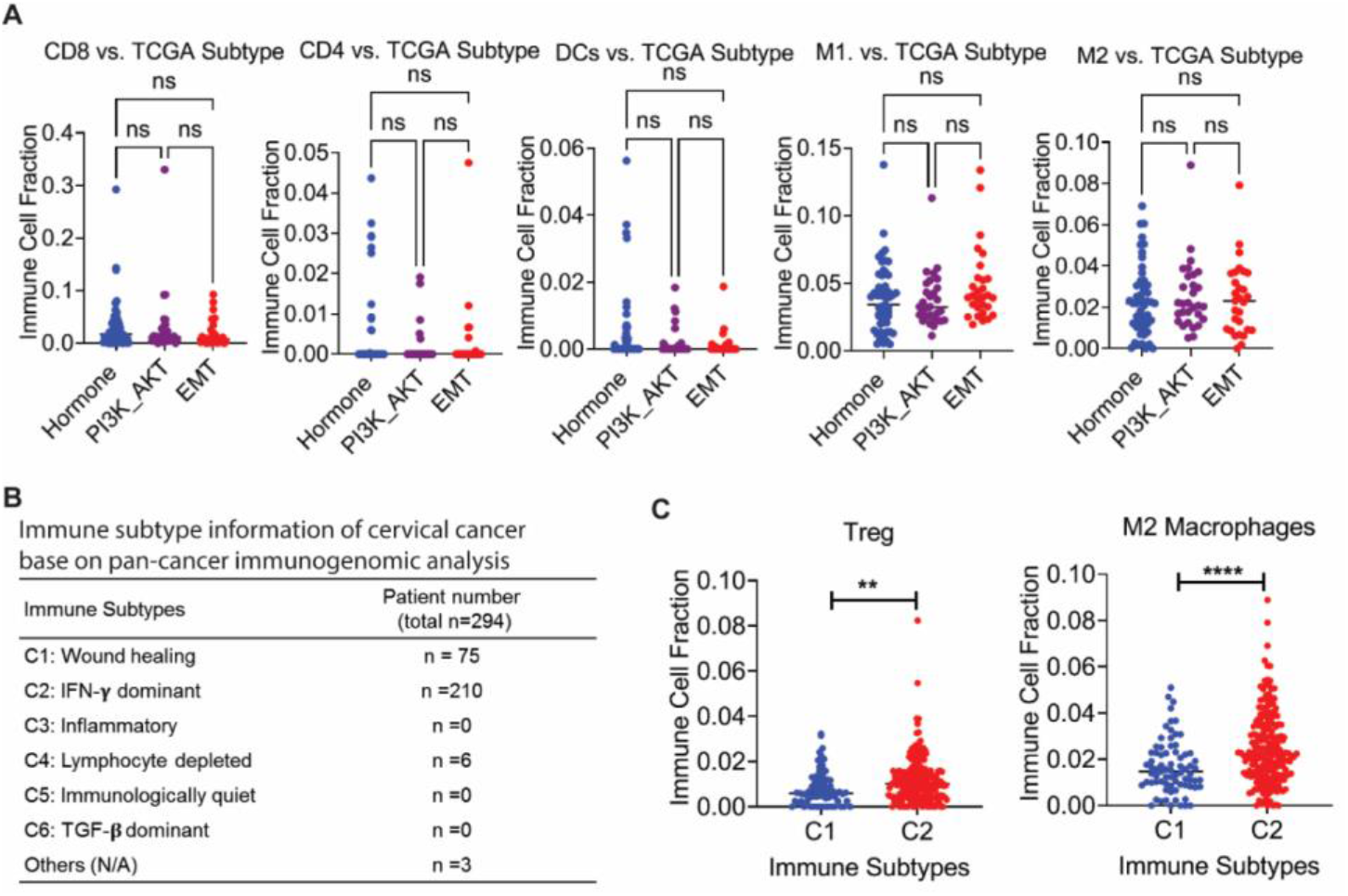
Immune cell infiltration in TCGA genetic subtypes and immunogenomic subtypes of cervical cancer. (A) Analysis of immune infiltrates immune cell fraction between three TCGA subtypes: Hormone, PI3K-AKT, and EMT. ANOVA with Tukey’s post-test was too used to analyze differences. P values < 0.05 were considered statistically significant. (B)& (C) Analysis of immune infiltrated immune cell fraction between immunogenomic subtypes: C1 and C2. T-test was used to analyze differences between two subtypes. P-values < 0.05 were considered statistically significant.

A recent study conducted by Thorsson et. al. identified six immune subtypes based on a pan-cancer immunogenomic analysis across 33 types of cancers [10]. We found that most cervical cancers were clustered in two of six immunogenomic subtypes (Table 1), subtype C1 (wound healing) and C2 (IFN-ɤ dominant). Through T-test with unequal variances, we found that C2 patients contained a significantly higher concentration of Regulatory T cells and M2 macrophages (Figure 6B). This significant difference in terms of immune landscape between the C1 and C2 cervical cancer patients can serve as implications for personalized treatment strategies or the increase in usage of different treatment strategies for different immunogenomic patient groups in order to account for the significant differences in immune landscape and create a better prognosis.

## 4. Discussion

Immunotherapies have experienced profound responses and unprecedented clinical benefits in many cancers, including previously terminal stages of relapsed or refractory B-cell lymphoma, advanced non-small-cell lung cancer (NSCLC), and metastatic melanoma [5, 12, 24, 32-36]. Unfortunately, the same treatment only showed modest clinical benefits in a small subset of patients with cervical cancers [8]. Results of clinical trials and preclinical investigations suggest that there are potently immunosuppressive factors that predominate the microenvironment and inhibit anti-tumor immunity in cervical tumors. Therefore, understanding the interactions among the tumor cells, immune cells, and other components of the tumor microenvironment may provide ways to improve the development of the next generation of immunotherapies. In this study, we have presented a comprehensive overview of the immune cell environment in cervical cancer. We characterized cellular compositions of immune infiltrates of cervical cancers compared the immune infiltrates of cervical cancer with other cancers. We also analyzed the correlation between immunogenetic phenotype and patient prognosis. By comparing HPV positive and negative head and neck cancer, we explored the associations between tumor immune infiltrates and HPV infection. Overall, this study provides a conceptual framework to understand the tumor immune microenvironment of cervical cancer. The major limitation of our study is the lack of bench work-based data support. In the future, other methods such as FACS-based immune profiling with direct patients’ samples would make the conclusion more convincing.

A ‘pro-inflammatory’ phenotype, such as increased infiltrating T lymphocytes, was demonstrated associated with improved clinical outcomes in many cancers [9, 12, 26, 27, 37-39]. Cervical cancer had a relatively higher average level of infiltrated CD8 T cells than other cancer (Figure 2). Moreover, tumor infiltrated CD8 T cells was favorable prognostic marker in cervical cancer (Figure 4). These results suggesting that CD8 T cells play a critical role in controlling cancer progression and a combination of standard ICB treatments with cytokines for inducing CD8 T cell enrichment may lead to improved clinical outcomes in cervical cancer. An ‘immunosuppressive’ microenvironment contains a high fraction of inhibitory immune cells, such as M2-like macrophages and regulatory T cells. An ‘immunosuppressive’ phenotype is usually considered as an unfavorable factor for cancer patients [10, 13, 21, 29, 40]. However, infiltrating M2-like macrophages was associated with a better prognosis in cervical cancer (Figure 4). In fact, a similar phenomenon was also observed in some other cancer cells, for example, patients with higher tumoral infiltration of Treg cells also had a better prognosis in pancreatic, colorectal, bladder, and esophageal cancers [41-43]. These results indicate that the tumor microenvironment has a complex cellular cross-talking. A recent study reported that Treg could reprogram the fibroblast population and restrict tumor progression in pancreatic cancer [43]. Therefore, the role of immunosuppressive M2 macrophages in cervical cancer progression, as well as in the response of cancer treatments, needs to be further investigated.

The immune system uses innate and adaptive immunity to recognize and combat foreign agents that invade the body, but such an immune response is usually not demonstrable or insufficient against HPV and all cervical cancer patients were HPV positive [44, 45]. However, our results showed that HPV-positive tumors had more infiltrated CD8 T cells than HPV-negative tumors, indicating that HPV still induces a certain degree of immune response and HPV-based therapeutic cancer vaccines have potential clinical benefit in the cervical tumors. Indeed, monotherapy of prophylactic L1 virus-like protein vaccines for HPV 16 had effectively induced durable regressions of premalignant HPV16–induced anogenital lesions [46, 47]. More importantly, in a recent phase I-II study, the combination of an HPV-targeting vaccine and standard-of-care chemotherapy were used to treat patients with advanced, recurrent, or metastatic cervical cancer, resulted in regressions and stable disease in more than 40% (32 of 72) of evaluable patients. The group of patients with higher vaccine-induced T cell immunity had a longer survival time with a flat tail on the survival curve [48]. These findings demonstrate that HPV-based vaccination can be exploited to combine with other therapies and benefit patients with advanced cervical cancer.

## 5. Conclusions

In summary, we analyzed ten infiltrated immune cells and provided a comprehensive landscape of the immune micro-environment in cervical cancers. We determined the prognostic value of a specific subtype of tumor-infiltrating immune cells for clinical outcomes in cervical cancers. Our results showed that cervical cancer shared a similar immune infiltrated microenvironment with other squamous cell carcinomas and CD8 T cell infiltration was associated with a better prognosis in cervical cancer. We also demonstrated that HPV infection affected immune cell infiltration and induce pro-inflammatory phenotypes. Our study provides a systematic insight into the tumor immune microenvironment of cervical cancers and offers a conceptual framework for the future design of rational combination treatment strategies to improve immuno-therapy outcomes.

## Data Availability

The data presented in this study are openly available in the TCGA database. https://portal.gdc.cancer.gov/legacy-archive/search/f

## Author Contributions

Xinyue Fan contributed to data collection, data analysis, and manuscript preparation. Chunbo He supervised these studies and contributed to the study design, data analysis, and manuscript preparation. All authors have read and agreed to the published version of the manuscript.

## Funding

This work was supported by the Fred & Pamela Buffett Cancer Center.

## Institutional Review Board Statement

Ethical review and approval were waived for this study, due to all data were from existing and de-identified public datasets. The TCGA Publication Committee ensured that our data’s status is categorized as “No restrictions; all data available without limitations.”

## Informed Consent Statement

Informed patient consent was waived for all data we used due to all data were from existing and de-identified public datasets. The TCGA Publication Committee ensured that our data’s status is categorized as “No restrictions; all data available without limitations.”

## Data Availability Statement

The data presented in this study are openly available in TCGA database. The raw data, processed data, and clinical data can be found in the legacy archive of the National Cancer Institute’s Genomic Data Commons (GDC) (https://portal.gdc.cancer.gov/legacy-archive/search/f).

## Conflicts of Interest

The authors declare no conflict of interest.

## Notes

### Competing Interest Statement

The authors have declared no competing interest.

### Author Declarations

Ethical review and approval were waived for this study, all data were from existing and deidentified public datasets.

